# Digital tools to improve parenting behavior in low-income settings: A mixed-methods feasibility study

**DOI:** 10.1101/2022.10.31.22281757

**Authors:** Lena Jäggi, Leonel Aguilar Melgar, Milagros Alvarado Llatance, Andreana Castellanos, Günther Fink, Kristen Hinckley, Maria-Luisa Huaylinos Bustamante, Dana C. McCoy, Hector Verastegui, Daniel Mäusezahl, Stella M. Hartinger Pena

**Author notes:** Corresponding author Lena Jäggi, lena, **Swiss Tropical and Public Health Institute**, Kreuzstrasse 2, 4123 Allschwil, Switzerland. joint senior authors (Daniel Mäusezahl, Stella M. Hartinger Pena). Financial disclosure: Andreana Castellanos is the CEO of Afinidata. All other authors have no financial interests to disclose.

## Abstract

**Introduction:** Digital parenting interventions could be potentially cost-effective means for providing early child development services in low-income settings. This 5-month mixed-methods pilot study evaluated the feasibility of using *Afinidata*, a comprehensive *FBMessenger*-based digital parenting intervention in a remote rural setting in Latin America and explored necessary adaptations to local context.

**Methods:** The study was conducted in three provinces in the Cajamarca region, Peru, from February to July 2021. 180 mothers with children aged between 2-24 months and regular access to a smartphone were enrolled. Mothers were interviewed three times in-person. Selected mothers also participated in focus groups or in-depth qualitative interviews.

**Results:** Despite the rural and remote study site, 88% of local families with children between 0-24 months had access to internet and smartphones. Two months after baseline, 84% of mothers reported using the platform at least once, and of those, 87% rated it as useful to very useful. After 5 months, 42% of mothers were still active on the platform, with little variation between urban and rural settings. Modifications to the intervention focused on assisting mothers in navigating the platform independently and included adding a laminated booklet with general information on child development, sample activities and detailed instructions on how to self-enroll in case of lost phones.

**Conclusions:** We found high access to smartphones and the intervention was well received and used in very remote areas of Peru, suggesting that digital parenting interventions could be a promising path forward for supporting low-income families in remote parts of Latin America.

- **What is already known on this topic** – *Digital parenting interventions are potentially cost-effective means for providing early child development services in low-income settings but have not been well-studied to date*.
- **What this study adds** – *This mixed-methods pilot study in a very remote setting in Latin America showed that 88% of local families with children between 0-24 months had access to smartphones with internet and 42% were still engaging with a digital parenting intervention after 5 months*.
- **How this study might affect research, practice or policy** – *High access to smartphones suggest that digital parenting interventions could be a promising path forward for supporting low-income families in remote parts of Latin America, but mothers might benefit from additional instructions to navigate all features of digital interventions on their own*.

## OBJECTIVE

According to latest estimates, over 250 million young children are at risk of missing their developmental potential due to early life adversity.[1] Early life interventions are increasingly recognized as key for ensuring children’s optimal development and long-term well-being.[2,3]

One of the most promising interventions for low-income settings are home visiting programs, where trained facilitators regularly visit parents following a structured curriculum to improve their knowledge and care practices related to early child development (ECD).[4,5] Such programs have been successfully implemented across the world[6–9] and in Peru.[10] The national Peruvian home visiting program (*Programa Nacional Cuna Más*, PNCM[11]) is one of the world’s largest, serving families with children under 36 months of age from areas of poverty. While a recent evaluation has shown PNCM to positively impact ECD and be cost-effective at scale[12], the program can currently only support the most vulnerable.

The rapid rise in cell-phone coverage and internet access globally means digital parenting interventions may be one means for addressing gaps in ECD services in Peru and many other low- and middle-income country (LMIC) settings.[13,14] *Afinidata* is one example of a comprehensive digital parenting support platform leveraging this opportunity. Intended for children under six, *Afinidata* uses dynamically collected user feedback to provide parents with customized activity recommendations that support ECD and learning using a *Facebook (FB) Messenger* chatbot.[15] *Afinidata* was designed focusing specifically on the needs of LMIC families and includes minimal text and data-intensive features.

Despite their considerable potential, there is limited evidence on how feasible digital parenting interventions are in practice, especially in a remote rural setting in a LMIC context. The current study describes the results of a 5-month mixed-methods study testing reach, reception, and use of the *Afinidata* parenting platform while exploring necessary adaptations to local context. We also summarize lessons learned for the implementation of *Afinidata* in the subsequent full-scale RCT (https://clinicaltrials.gov/ct2/show/NCT05202106).

## METHODS

### Design and Ethical Approval

The results reported in this feasibility pilot study follows the framework for feasibility studies[16] and STROBE standards.[17] The study was approved by the Universidad Peruana Cayetano Heredia (SIDISI: 202522-Ref 030-03-21) and the Ethics Commission for Northwest and Central Switzerland (EKNZ: AO_2021-00002). Community leaders and local authorities were aware of the study.

### Setting

The study was conducted in the provinces of San Marcos, Cajabamba and Cajamarca in northern Andean Peru, located between 1900 and 3900 meters above sea level. The region is predominantly rural and representative of many rural and peri-urban settings in Andean South America, with a large share of poor and remote households engaged in farming. Communities were classified in accordance with the Peruvian National Institute of Statistics and Informatics (INEI) as urban (communities with more than 2,000 inhabitants with contiguous, grouped homes forming streets) or rural (scattered or grouped houses of up to 2,000 inhabitants per community[18]). The study area is partially covered by PNCM.

### Participants

We recruited 180 adult mothers with children aged between 2-24 months who either owned or had regular access to a smartphone and who were living across 49 communities representative for the three provinces.

### The Intervention

The *Afinidata* platform (www.afinidata.com) uses *FBMessenger* to interact with pregnant mothers and parents of children aged 0-6 years from low-income settings through automated chatbots. Similar to home visitors, a “virtual tutor” asks about the child’s well-being through messages and push-notifications and makes 2-3 suggestions for development-promoting activities that parents can do with their children. Parents can reach out to the chatbot anytime, seeking advice or sharing their child’s achievements. The system tracks parents’ reactions to the recommended activities and children’s development over time.

During the study, mothers received a weekly push-message with age-appropriate informative content and a reminder to use *Afinidata*. Mothers could also respond to questions about their children’s development, which were summarized into a simple graphical report. We screened all 641 activities and messages for the relevant age group (2-24 months) for local language, cultural appropriateness and availability of playing materials.

### Procedures

We conducted five months of extensive field-testing with local mothers from February 2021 to July 2021, squarely during the COVID-19 pandemic in Peru.

First, we measured internet and phone connectivity in 142 communities in all three provinces for each of the four national cell-phone providers. Connectivity measurements were taken in the central plaza of each community, or in central schools, health centers, or main roads. Using SIM cards for each provider in the same ZTE-BLADE-A3-2020 smartphone, we started a conversation with *Afinidata* in *FBMessenger* and opened the platform-generated link for an activity. Of the 136 communities with successful activity-download through at least one of the four providers, we selected 49 communities for this study. We also noted presence of administration building, health center and school for each community. Using administrative data on recent births, we identified all eligible families and visited them at home. Participating families signed an informed consent. No incentives were given for participation. Mothers were shown how to use *Afinidata* and completed a baseline interview. After two months, mothers gave detailed qualitative and quantitative feedback on *Afinidata* content during a second in-person visit. Simultaneously, we called 27 mothers who were not using *Afinidata* to inquire about technical difficulties and reasons for non-use and conducted seven focus groups and two in-depth interviews with a mix of urban and rural mothers with varying levels of engagement in *Afinidata*. After five months we conducted a final in-person visit asking about additional characteristics of the family, mothers’ digital literacy, and their platform use and satisfaction.

### Measures

#### Main outcome measures

Use of the app over time was measured automatically by *Afinidata*. We defined mothers as active users if they interacted with *Afinidata* at least once per week, either asking for an activity, choosing a *Frequently Asked Question* or updating their child’s development. Some other interactions with the platform, such as responding to push-messages or reminders, requesting and receiving tips on development, or asking direct questions to the virtual tutor, were not tracked in the system.

#### Demographic, socioeconomic and literacy measures

Sociodemographic data were collected at baseline and included household characteristics, reception of conditional cash transfers for poor families called “Juntos”,[19] mother’s and child’s age (in years and in months, respectively), mother’s education and economic activity. For reading comprehension, mothers answered three content questions after reading a sample activity. Results are presented as percentage who answered all questions correctly.

#### Internet connectivity, phone use and digital literacy

At baseline, mothers reported internet signal availability in the house, their phone ownership, type of contract, monthly cost and social media usage. At endline, mothers also reported on their satisfaction and future use of *Afinidata* and completed the Survey of Adult Skills digital literacy scale (PIAAC; Cronbach’s alpha=0.79).[20]

### Data analysis

Statistical analyses were conducted using STATA16 and Python’s statsmodels and scipy libraries. Participant characteristics are presented as means, standard deviations and relative percentages. We compared participants from urban and rural communities mothers who were active versus inactive at endline using chi-square and two sided Mann-Whitney U rank tests (non-corrected for continuity). Additionally, we analyzed use over time for all participants by mapping the percentage of mothers who were active in a given week by phone ownership and setting (urban/rural).

In the focus groups and qualitative interviews we asked in-depth questions on *Afinidata* satisfaction and specific in-platform features. All transcripts and notes were summarized and analyzed for common themes.

## RESULTS

### Reach of the Digital Intervention

The five urban communities included in the study had full internet connectivity for all providers and all resources present. Among the 44 rural communities, 10 (22%) had connectivity with only one or two phone providers. A participant flowchart is included in Figure 1 and after up to three attempts at contact, 133 mothers were interviewed at a final visit. A general description of the sample, as well as phone and social media use by urban and rural setting, is included in Table 1. Of all eligible families, 12% (35) were excluded because there was no smartphone available for the mother to use. Among included families, 16% (28) of mothers used a phone belonging to their husband or another family member rather than their own.

**Table 1.** Demographic Characteristics of Sample.

|  | Total |  | Urban |  | Rural |  | Diff. Test |
| --- | --- | --- | --- | --- | --- | --- | --- |
|  | N/Mean | Column %/SD | N/Mean | Column %/SD | N/Mean | Column %/SD |  |
| Participants (N, row %) | 180 | 100.0% | 41 | 22.8% | 139 | 77.2% |  |
| **Family Characteristics** |  |  |  |  |  |  |  |
| Age child (months) | 9.63 | 4.84 | 10.75 | 5.7 | 9.3 | 4.53 | n.s. |
| Age mother (N=179, years) | 29.08 | 6.13 | 28.77 | 6.08 | 29.17 | 6.17 | n.s. |
| Education mother (completed secondary or higher, N=178) | 56 | 31.5% | 21 | 51.2% | 35 | 25.5% | ** |
| Mother is mainly at home with children (*Ama de casa*) | 153 | 85.0% | 29 | 70.7% | 124 | 89.2% | ** |
| Mother answered all reading comprehension questions correctly | 143 | 79.4% | 37 | 90.2% | 106 | 76.3% | n.s. |
| Agriculture is main economic activity in household (N=133) | 69 | 51.9% | 10 | 32.3% | 59 | 57.8% | * |
| House has dirt floor | 69 | 38.3% | 2 | 4.9% | 67 | 48.2% | *** |
| Household received conditional cash transfers | 98 | 54.4% | 18 | 43.9% | 80 | 57.5% | n.s. |
| **Access to community resources** |  |  |  |  |  |  |  |
| School in the community | 176 | 97.8% | 41 | 100.0% | 135 | 97.1% | n.s. |
| Health center in the community | 106 | 58.9% | 41 | 100.0% | 65 | 46.8% | *** |
| Local administration building in community | 172 | 95.6% | 41 | 100.0% | 131 | 94.2% | n.s. |
| **Internet connectivity and digital literacy** |  |  |  |  |  |  |  |
| Number of providers in community | 3.77 | 0.56 | 4.00 | 0.00 | 3.70 | 0.62 | *** |
| Household has internet signal in the house | 169 | 93.9% | 40 | 97.6% | 129 | 92.8% | n.s. |
| Mother owns smartphone | 152 | 84.4% | 39 | 95.1% | 113 | 81.2% | * |
| Prepaid phone (vs. monthly plan) | 119 | 66.1% | 22 | 53.7% | 97 | 69.8% | n.s. |
| Monthly cost of phone (Soles): |  |  |  |  |  |  | * |
| less than 16 Soles (4 USD) | 67 | 37.2% | 11 | 26.8% | 56 | 40.3% |  |
| between 16-30 Soles (4-8 USD) | 90 | 50.0% | 20 | 48.8% | 70 | 50.4% |  |
| over 30 Soles (8 USD) | 23 | 12.8% | 10 | 24.4% | 13 | 9.4% |  |
| Digital Literacy Score (N=133) | 10.86 | 4.5 | 14.35 | 4.54 | 9.8 | 3.93 | *** |
| **Social media usage (self-report)** |  |  |  |  |  |  |  |
| Mother uses *FB* (N=179) | 169 | 94.4% | 41 | 100.0% | 128 | 92.8% | n.s. |
| Mother uses *FB Messenger* (N=179) | 152 | 84.9% | 40 | 97.6% | 112 | 81.2% | ** |
| Mother uses *Whatsapp* (N=179) | 173 | 96.6% | 41 | 100.0% | 132 | 95.7% | n.s. |
| ***Afinidata* usage (self-report after 5 months)** |  |  |  |  |  |  |  |
| Mother used *Afinidata* (N=133) | 128 | 96.2% | 30 | 96.8% | 98 | 96.1% | n.s. |
| Mother rated *Afinidata* positively (Useful or Very useful, N=128) | 123 | 96.1% | 30 | 100.0% | 93 | 94.9% | n.s. |
| Mother intends to use *Afinidata* in the future (N=128) | 126 | 98.4% | 30 | 100.0% | 96 | 98.0% | n.s. |
| *Note.* FB= Facebook, n.s.= non-significant *p*-value. SD = Standard Deviation<br>*\*p*=05. *\*\*p*=01. *\*\*\*p*=001. |  |  |  |  |  |  |  |

**Figure 1.**
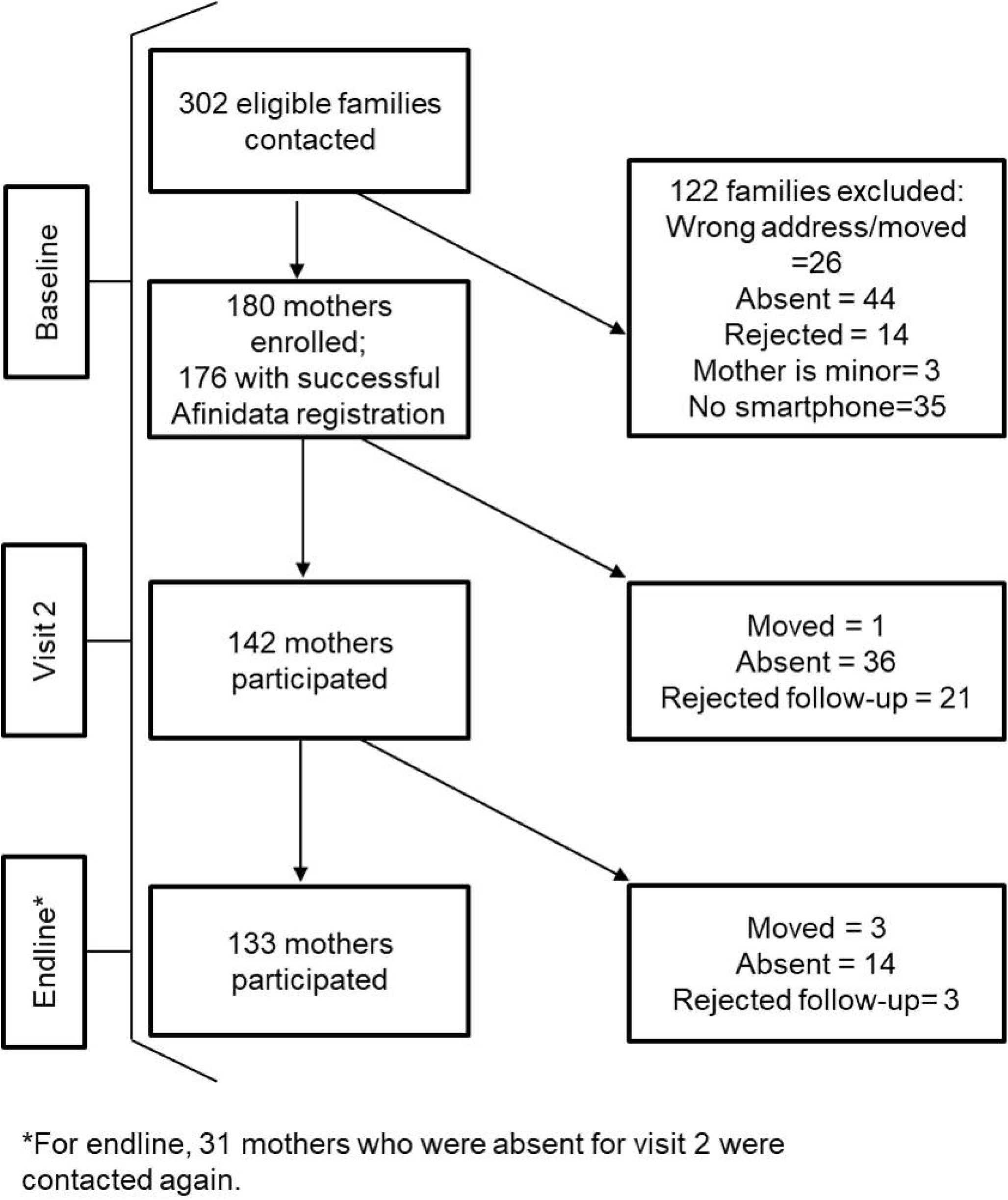
Flowchart of study enrollment

### Reception and Use of the Intervention

Two months after baseline, 84% of all mothers reported ever using the platform after the baseline visit and of those, 87% rated it as useful to very useful (mean=4.37/5, SD=1.00). These results are similar to automatically collected usage rates by the *Afinidata* platform after one month (see Figure 2a). Figure 2a also presents weekly usage rates collected by the *Afinidata* system for mothers who had their own smartphone and those who used a family member’s phone, and shows generally higher engagement by mothers with their own phones. Figure 2b presents the same analysis by urban versus rural setting, and shows no meaningful differences based on context. Overall, 42% (73) of mothers were still active after 5 months. These mothers did not differ by age of the child (under or over 12 months), mother’s level of education or social media usage at baseline.

**Figure 2a.**
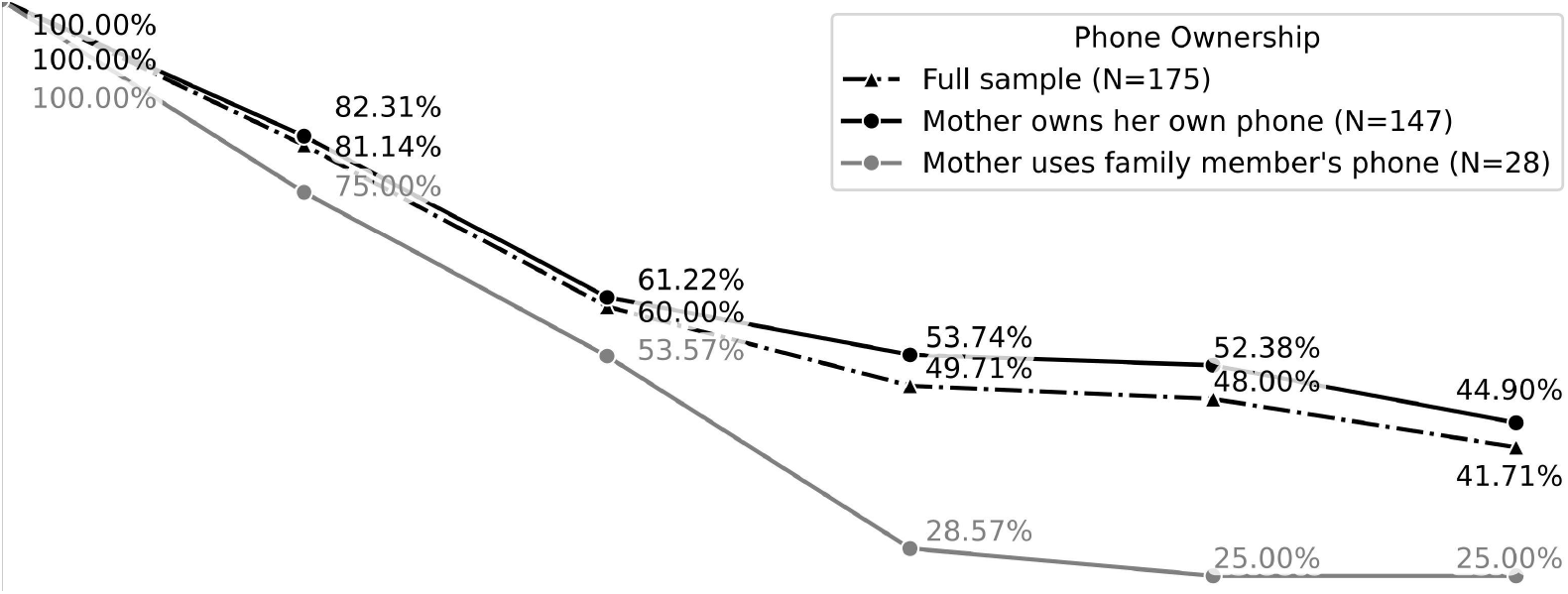
Afinidata platform use over time by phone ownership.

**Figure 2b.**
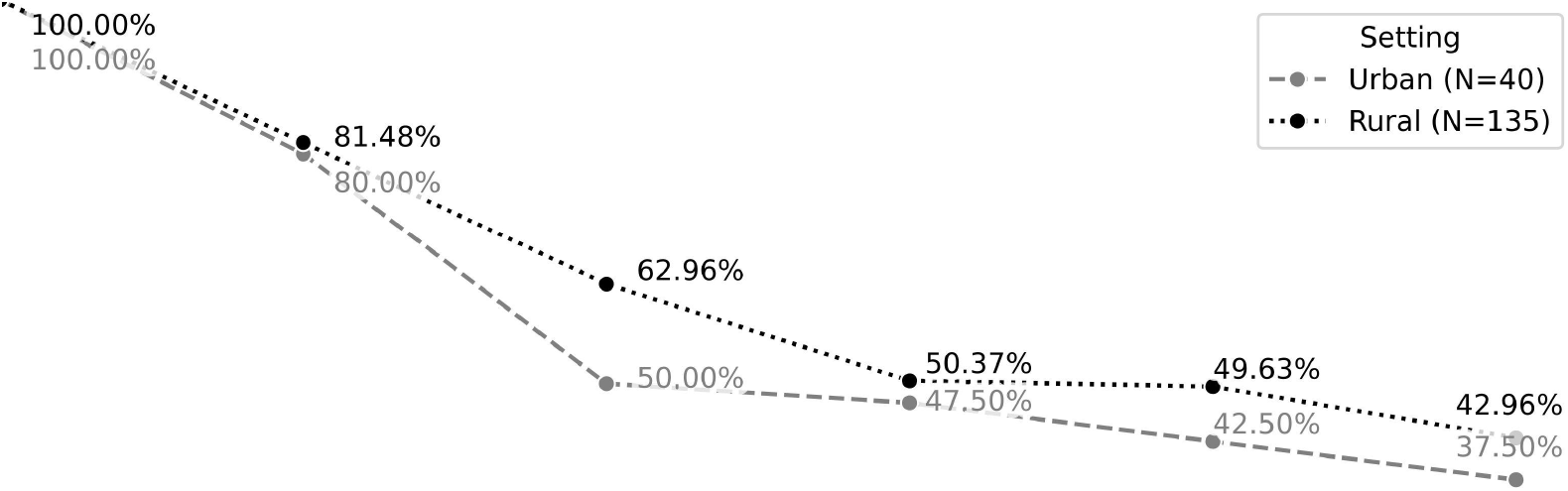
*Afinidata* platform use over time by urban and rural setting.

During the second visit, mothers also gave feedback on specific activities they had received. Most mothers reported spending between 20-30 minutes on the activities per session. Only 11% of mothers recalled an activity they did not like. Recommendations for improvement included simpler wording and materials. Interviews with the 16% of mothers who self-reported not using the platform revealed that barriers were mainly lack of time, followed by lost phone or lost access to a borrowed phone. Similarly, follow-up calls to 27 mothers without activity in the platform for over a month showed that mothers often did not re-install the platform if they switched phones.

In five focus groups and two in-depth interviews mothers expanded on the topics above. Overall, *Afinidata* was received very positively, and several mothers recounted spending more time engaging with their children, involving their husbands in the activities and feeling closer to their children. Mothers who experienced problems with materials reported substituting or moving on to the next activity without this having a negative impact on their perception of the intervention. Most mothers cited time constraints as the main barrier for participation. For those mothers with school-aged children, this was mainly due to additional demands on managing virtual classes during the COVID-19 pandemic. Additionally, sometimes mothers lost access to their phone because their older children needed it for school. Technical difficulties, lack of credit or loss of phone were not perceived as relevant barriers for use.

### Adaptation of the Intervention

We changed various components of *Afinidata* or the RCT protocol following the detailed feedback (see Table 2 for a summary). For example, to facilitate self- or re-enrollment by the mother, we designed a laminated booklet containing general information on ECD, sample activities and detailed self-enrollment instructions. To increase comfort and agency in using the platform, interviewers assist mothers in experiencing the platform features during the baseline visit of the RCT, and we added real-time communication between the field team and *Afinidata* to provide technical support during enrollment.

**Table 2.** Summary of Modifications to the Intervention Based on Participant Feedback

| Issue | Data source | Adaptation to Intervention or Study Protocol |
| --- | --- | --- |
| **Optimizing procedures for subsequent trial** |  |  |
| Field team and *Afinidata* need real-time communication channels to troubleshoot enrollment issues | Field team observations | • **Designated *Whatsapp* group between field team and *Afinidata* back-end team for technical support and to ask for manual data correction or enrolment** |
|  |  | • Field team has access to *Afinidata* dashboard<br>• Development of detailed trainings and technical manuals on how to trouble-shoot enrolment problems with the family |
| Mothers have problems re-enrolling into *Afinidata* after loss of phone:<br>Several mothers self-registered with another FB account, generating multiple ID's under different names | Field team observations, user surveys, focus groups | • Adapted enrolment protocol and developed in-depth training manuals for field interviewers: Field workers spend more time to assist mothers in self-enrolment and give instructions on all available options at baseline visit. Balance between accurate data entry and letting mothers experience the interaction with *Afinidata* on their own to increase knowledge and comfort with the chatbot.<br>• *Afinidata* incorporated back-end programming to associate users who re-register with previous accounts<br>• Field workers leave a printed booklet with sample activities and detailed information on how to re-enroll in case mothers switch phones |
| 12% of approached households do not have access to a smartphone | User surveys, field team observations | • Field workers leave a printed booklet with general information on child development, sample activities and detailed information on how to self-enroll when a smartphone is available at a later point in time |
| Find additional strategies to increase retention | Compliance data, user surveys | • Protocol and scripted calls every 3 months to inactive users to inquire about technical difficulties. |
| Additional support to resolve technical difficulties during sign-up is needed | Field team observations | • Adding a new technical support button to the platform that connects directly to an *Afinidata* collaborator<br>• Field workers are showing mothers how to ask for technical support within the chatbot at baseline visit |
| Some mothers are asking interviewers for help on how to disengage from the program.<br>FB blocked the chatbot for all mothers for 48 hours after mothers blocked the *Afindata* chatbot | Field team observations, "FB block" | • Included programming for users to drop out directly within the platform to avoid users blocking messages<br>• During enrollment field interviewers explain what to do if parents prefer to stop receiving messages and content from *Afinidata*<br>• Developed an additional application to secure alternative means of communication in case of a "FB block" as an alternative channel of communication |
| **Adaptation to local context** |  |  |
| Photos and descriptions of certain activities were not representative of local cultural context or were | Field team observations | • New photos were taken for 114 activities<br>• *Afinidata* made 214 adaptations to content such as replacing words or substitute |
| depicting toys or resources not readily available in the local cultural context |  | **materials that were costly or hard to find locally**<br>• **Afinidata categorized all existing content based on the Nurturing Care Framework to assist in a more balanced coverage of topics around health, nutrition, responsive caregiving and other topics.** |
| Limit amount of questions needed to generate information on developmental milestones | Focus groups | • **Development of a new algorithm to place a child in the appropriate *Afinidata* developmental status groups** |
| Make report on developmental milestones more understandable for families | Focus groups | • **Testing of different layout options and final development of a new report on developmental progress of the child for the families.** |
| Optimize push-message timing and frequency | Focus groups | • **Changed time of day when push notifications are sent to users** |

## DISCUSSION

Digital parenting interventions are one potentially cost-effective yet untested means for expanding access to ECD programs in LMICs. In this study, we extensively tested the reach, reception, and use of *Afinidata*, a *FBMessenger*-based “virtual tutor” for parents, and explored necessary adaptations to local context in a remote rural setting in Latin America.

Our results show that there was high access to internet and smartphones throughout the rural and remote region. Only six of the most remote villages out of 142 communities were excluded from the study area for lack of internet connectivity. On an individual level, 88% of the eligible families had a smartphone and 94% of participating households had internet signal inside their homes. Moreover, 84% of mothers owned their own device, and almost all were already familiar with and using *Facebook* (94%) and *Whatsapp* (96%). This shows a tremendous potential to reach most of the primary caregivers of young children through interventions that take advantage of those channels in a typical LMIC setting.

In terms of content, the *Afinidata* platform was received very positively by mothers in our study. A high proportion of mothers visited at endline reported that they would continue to use *Afinidata* in the future. However, this contrasts with actual usage data from the platform. While there was high initial engagement with *Afinidata*, there was a rapid decline in use over two months, and after five months only 42% (73) of mothers were still engaging weekly. Digital parenting interventions commonly have to contend with high attrition rates.[21,22] In programs that resemble *Afinidata* (i.e. freely available online-only universal prevention programs for parents), completion rates as low as 15%[23] to 7%[24] have been reported. Since we only measured part of the possible interactions with the system, this likely is an underestimation of engagement. Keeping users engaged through push-messages and other content is an important part to maintain interest in the intervention over time. The following RCT will measure these interactions and investigate additional barriers for engagement.

As expected, mothers who did not own their own phone showed lower rates of engagement than those who did, with only 25% still engaging weekly with the platform after 5 months. Importantly, however, we found no meaningful differences in engagement between urban or rural participants. Furthermore, there were no differences in use by mothers’ level of education, social media use or age of the child, indicating that the intervention is relevant and accessible for mothers across different contexts. Lack of credit, technical difficulties and loss of phone were not perceived as barriers for engagement by the mothers, though field observations showed that mothers often did not re-install the platform after they switched phones.

Because we found few concerns regarding the content of *Afinidata*, we focused on modifications that would assist mothers in navigating the platform independently in our RCT (e.g., teaching how to self-enroll, ask for technical support, or leave the platform). Two key lessons learned were: (1) While the use of the platform quickly becomes intuitive, mothers in remote areas with poor connection greatly benefit from personal assistance with the initial installation and exploration of the features. (2) It is beneficial to accompany the digital intervention with a physical booklet giving instructions for later self-enrollment for families who might not have a smartphone present during the visit or who change phones later.

## Limitations

This study had several limitations. Most importantly, we did not assess impact of the intervention on maternal or child outcomes – this will be done in the ongoing RCT. Our results could be biased by losing the least satisfied users for follow-up. While we tried to reach a sample typical for the region, excluding families without access to a smartphone means that this was not a truly representative sample.

## Conclusion

Our results show that access to smartphones is high in very remote areas of Peru, and that digital ECD interventions are well-received by mothers across urban and rural settings with diverse individual characteristics. Digital parenting interventions could thus be a promising path forward for supporting low-income families in remote parts of Latin America and other LMICs.

## Data Availability

All data produced in the present study are available upon reasonable request to the authors

## ACKNOWLEDGMENTS

We would like to thank our field staff for their excellent work and all their efforts, as well as study participants for their willingness to help us with this project.

